# A Practical Risk-Score for Early Prediction of Neurological Outcome after Out-of-Hospital Cardiac Arrest –MIRA_2_CLE_2_

**DOI:** 10.1101/2020.04.12.20045146

**Authors:** Nilesh Pareek, Peter Kordis, Nicholas Beckley-Hoelscher, Dominic Pimenta, Spela Tadel Kocjancic, Anja Jazbec, Joanne Nevett, Rachael Fothergill, Sundeep Kalra, Tim Lockie, Ajay M. Shah, Jonathan Byrne, Marko Noc, Philip MacCarthy

**Author notes:** **<u>Correspondence:</u>** Nilesh Pareek, School of Cardiovascular Medicine and Sciences, BHF Centre of Excellence, London. **Institution where work performed:** King’s College Hospital NHS Foundation trust, London, U.K. **First author:** Nilesh Pareek.

## Abstract

**Aims:** The purpose of this study was to develop a practical risk−score to predict poor neurological outcome after out−of−hospital cardiac arrest (OOHCA) for use on arrival to a Heart Attack Centre.

**Methods and Results:** Between May 2012 and December 2017, 1055 patients had OOHCA in our region, of whom 373 patients were included in the King's Out of Hospital Cardiac Arrest Registry (KOCAR). We performed prediction modelling with multi-variable logistic regression to identify factors independently predictive of the primary outcome in order to derive a risk score. This was externally validated in two independent cohorts comprising 474 patients. The primary outcome was poor neurological function at 6−month follow−up (Cerebral Performance Category 3-−). Seven independent variables for prediction of outcome were identified: Missed (Unwitnessed) arrest, Initial non-shockable rhythm, non-Reactivity of pupils, Age, Changing intra-arrest rhythms, Low pH<;7.20 and Epinephrine administration. From these variables, the MIRA2CLE2 score was developed which had an AUC of 0.90 in the development and 0.85 and 0.89 in the validation cohorts. 3 risk groups of the MIRA2CLE2 were defined − Low risk (≤2−5.6% risk of poor outcome; Intermediate risk (3−4−55.4% of poor outcome) and high risk (≥5−92.3% risk of poor outcome). The risk-score performance was equivalent in a sub-group of patients referred for early angiography and revascularisation where appropriate.

**Conclusions:** The MIRA2CLE2 score is a practical risk score for early accurate prediction of poor neurological outcome after OOHCA, which has been developed for simplicity of use on admission to a Heart Attack Centre.

## INTRODUCTION

Out of Hospital Cardiac Arrest (OOHCA) occurs in over a quarter of a million patients a year globally and presents a major public health challenge (1). There is an increasing recognition of a primary cardiac aetiology in OOHCA with the presence of a culprit coronary lesion in 50-90% of patients (2-4). As a result, a significant proportion of OOHCA patients are taken directly to a Heart Attack Centre (HAC) for consideration of trans-thoracic echocardiography, early coronary angiography and mechanical circulatory support (MCS), such as percutaneous left ventricular assist devices and extra-corporeal membranous oxygenation. These approaches have the potential to improve survival after OOHCA but are invasive and costly and, despite their application, a majority of patients still sustain poor outcomes due to hypoxic brain injury incurred prior to admission (5). Furthermore, the clinical features of neurological injury are usually delayed until 72 hours after admission, by which point, much resource has already been expended (6).

Early predictors of outcome that would support emergency clinical decision making in the HAC are urgently required to prevent instigating costly and intensive resources in cases of futility, to inform family discussions and to guide clinical decision making. The OHCA (7), CAHP (8) and TTM risk tools (9) have been developed but these are presented as relatively complex normograms, thereby potentially limiting their routine use in emergency settings.

Accordingly, the purpose of this study was to develop a practical point-based risk score which can be applied to patients with OOHCA on arrival to a HAC to reflect long term prognosis and support clinical decision making.

## METHODS

### Study Setting

A standardised systematic protocol was established in 2012 in London whereby patients who have sustained OOHCA with return of spontaneous circulation (ROSC) and an ECG showing ST elevation are taken directly to a HAC. Patients without ST elevation were brought directly to the HAC if there was high suspicion of a cardiac aetiology or after exclusion of non-cardiac causes in the emergency department. King’s College Hospital is the main HAC in South East London, treating a population in excess of 1 million. On arrival, a decision to perform coronary angiography is made by the admitting Interventional Cardiologist and after treatment, patients are transferred to the Intensive Care Unit for ongoing supportive care.

### Study Population

We included patients all patients over the age of 18 years who presented with OOHCA and had ROSC in the community between 1^st^ May 2012 and 1^st^ May 2017. Inclusion criteria for the registry was all patients with ST elevation on ECG and for patients without ST elevation, if there was no evidence of a non-cardiac aetiology. Patients who died before arrival to our centre, evidence of an obvious non-cardiac cause of arrest (suicide, trauma, drowning, substance overdose), patients with confirmed intra-cerebral bleeding, prior neurological disability (Cerebral Performance Category (CPC) 3 or 4) or any survival limiting disease were excluded. Any patients with intact conscious status (defined as a GCS of 15/15) on arrival were also excluded from the final analysis. The study was performed according the principles of Declaration of Helsinki and received approval by the Research Ethics Committee and Health Research Authority.

### Data Collection

Data was collected using a dedicated database and patients were identified using the Utstein criteria (10, 11). We formed a data collaboration with the London Ambulance Service to ensure accurate collection of pre-hospital data, including time of cardiac arrest, initial rhythm, use of bystander cardio-pulmonary resuscitation (CPR) and time of ROSC. Medical records were collected and included index arterial blood gases and standard biochemical and haematological measures (within 30 minutes of arrival). We also collected baseline cardiovascular investigations including ECG, emergency echocardiography and coronary angiography where appropriate.

### Outcome

The primary end-point was poor neurological outcome, classified as CPC 3-5 (severe disability–death) at follow-up of 6 months (blinded analysis). Follow-up was tracked using the medical records and we approached all patients who were alive at the point of follow-up to perform a telephone interview. All patients that died before 6 months were classed as CPC 5 other than those that were CPC 1 - 2 before an unrelated cause of death - these patients remained in this class for the purposes of the primary end-point analysis.

### Statistical Analysis

We developed the risk score in accordance with the TRIPOD (Transparent Reporting of a multivariable prediction model for Individual Prognosis Or Diagnosis) methodology (12). In accordance with the guidelines, logistic regression was used over Cox regression analysis owing to a shorter follow-up duration (12). An exploratory univariate analysis was performed for selected candidate predictors. Based on immediate availability of variables and strongest statistical association with outcome (p<0.05), a smaller subset of predictor variables was entered into a multi-variable model. Simplified versions of the multivariable model were then used to develop simple additive risk scores based on influential cut-off points for continuous variables. A Receiver Operating Characteristic (ROC) curve for each risk score was plotted, with the Area Under the Curve (AUC) measuring the discrimination. Calibration slopes were used to measure the calibration of the scores rather than the Hosmer-Lemeshow goodness-of-fit test in accordance with current expert consensus (13). Missing predictor variables used in the multivariable model were investigated for associations with other variables, with appropriate multiple imputation methods used to handle missing values. An internal validation was performed using 1000 bootstrap iterations, generating bias-corrected and accelerated (BCa) confidence intervals. The risk scores were then calculated for patients in the validation cohorts, with the discrimination and calibration measured by the AUC and calibration slope respectively. Finally, a pre-specified analysis evaluating the performance of the risk score for patients undergoing early angiography was performed. A p <0.05 was considered statistically significant. All analyses were undertaken using R version 3.5.3.

## RESULTS

### Development Cohort

Between 1^st^ May 2012 and 31^st^ December 2017, 1055 patients suffered OOHCA in the South London area, of whom 251 failed to regain ROSC with the emergency medical services. From this cohort, 129 died before reaching our tertiary centre, 635 patients reached with ROSC, and 236 patients were deemed to have a non-cardiac aetiology for the cardiac arrest. After excluding conscious patients (defined as a GCS of 15/15) and those lost to follow-up (1 patient), 373 patients were included in the development registry (**Figure 1**).

**Figure 1.**
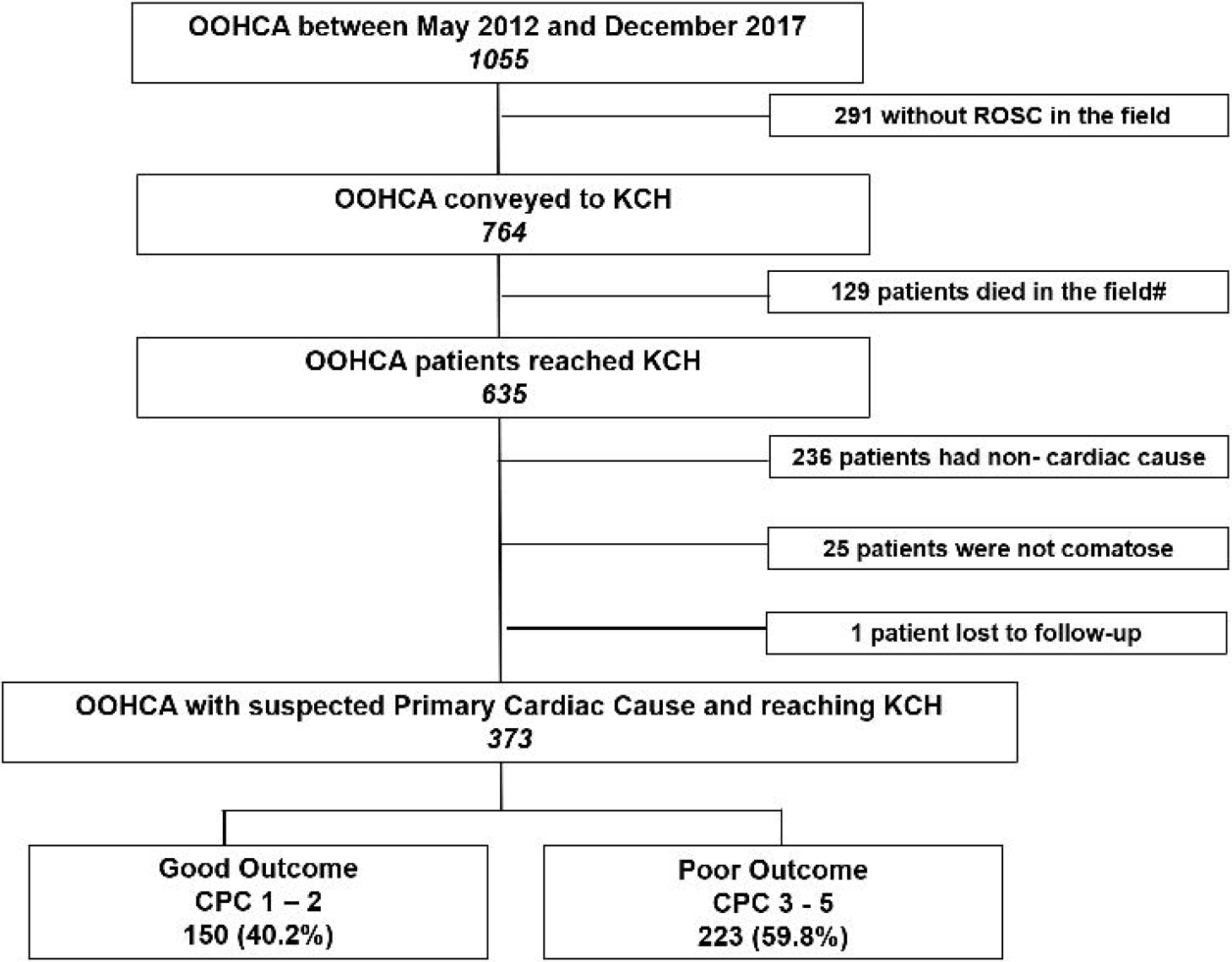
Flow chart of patient flow from the KOCAR registry cohort in the study.

Median age was 64.0 years, the majority of patients were male (74.2%), median zero flow time was 2 minutes (IQR 7) and low-flow time was 25 minutes (IQR 21). Most patients had cardiac arrest at home (n=212/373, 56.8%) and 263 patients (70.5%) had shockable rhythms. 209 patients had ST elevation/LBBB (56.1%) and 41 patients (11.0%) had ST depression on 12 lead ECG. Two hundred and sixty-two patients (70.2%) had early coronary angiography and 184 of these patients (70.2%) underwent percutaneous coronary intervention. The remaining 111 patients (29.8%) did not have coronary angiography based on clinician discretion. The primary endpoint of poor neurological outcome (CPC 3 – 5) occurred in 233 patients (59.8%) (**Tables 1 and 2**).

**Table 1.** Patient Baseline Characteristics.

| **Variables** | **Total (n=373)** | **Good Outcome**<br>**(n=150; 41.2%)** | **Poor Outcome**<br>**(n=223; 59.8%)** | **p** |
| --- | --- | --- | --- | --- |
| **Age (years)** | 64 (52 – 75) | 59 (49 - 68) | 68 (56 - 78) | <0.0001 |
| **Male - no./total no. (%)** | 277/373 (74.2) | 110/150 (73.3) | 167/223 (74.9) | 0.736 |
| **Cardio-vascular Risk factors - no./total no. (%)** |  |  |  |  |
| **Diabetes Mellitus** | 66/373 (17.6) | 13/150 (8.7) | 53/223 (23.8) | 0.002 |
| **Smoking** | 203/356 (57.0) | 82/149 (55.0) | 121/207 (58.5) | 0.520 |
| **ECG – no. (%)** |  |  |  |  |
| **ST elevation/**<br>**Left Bundle**<br>**Branch Block** | 209 (56.1) | 92 (66.0) | 117 (52.5) |  |
| **ST depression** | 41 (11.0) | 20 (13.3) | 21 (9.4) |  |
| **Right Bundle**<br>**Branch block** | 44 (11.8) | 7 (9.3) | 37 (17.0) |  |
| **Normal** | 78 (20.9) | 31 (20.7) | 47 (21.1) |  |
| **Left Ventricular Ejection**<br>**Fraction (%)** | 40 (30 - 50) | 45 (37.5 – 50) | 40 (30.0 – 50.0) | 0.067 |
| **Coronary Angiography -**<br>**no./total no. (%)** | 262/373 (70.2) | 125/150 (83.3) | 137/223 (61.4) | < 0.0001 |
| **PCI - no./total no. (%)** | 184/262 (70.2) | 95/125 (76.0) | 89/137 (64.9) | < 0.0001 |

**Table 2.** OOHCA Circumstances.

| **Variables** | **Total**<br>**(n=373)** | **Good Outcome**<br>**(n=150; 41/2%)** | **Poor Outcome**<br>**(n=223; 59.8%)** | **p** |
| --- | --- | --- | --- | --- |
| **Residence - no./total**<br>**no. (%)** | 212/373 (56.8) | 71/150 (47.3) | 141/223 (63.2) | 0.0001 |
| **Witnessed - no./total**<br>**no. (%)** | 293/372 (78.8) | 136/149 (91.3) | 157/223 (70.4) | <0.0001 |
| **Bystander CPR -**<br>**no./total no. (%)** | 266/371 (71.7) | 118/149 (79.2) | 148/222 (66.7) | <0.0001 |
| **Zero Flow Time**<br>**(mins)** | 2 (0 - 7) | 1 (0 - 4) | 3 (0 - 8) | 0.0001 |
| **Low Flow Time**<br>**(mins)** | 25 (17 – 38) | 19.0 (13.0 – 27.0) | 31 (21.0 – 43.0) | <0.0001 |
| **Shockable rhythm -**<br>**no./ total no. (%)** | 263/373 (70.5) | 141/150 (94.0) | 122/223 (54.7) | <0.0001 |
| **Reactive Pupils -**<br>**no./total no. (%)** | 144/321 (44.9) | 88/127 (69.3) | 56/194 (28.9) | <0.0001 |
| **Two Rhythms -**<br>**no./total no. (%)** | 152/374 (40.6) | 25/150 (16.7) | 127/224 (59.7) | <0.0001 |
| **Adrenaline - no./total**<br>**no. (%)** | 266/370 (71.9) | 63/149 (42.3) | 203/221 (91.9) | <0.0001 |
| **Admission pH** | 7.21 (7.08 – 7.30) | 7.27 (7.20 -7.33) | 7.17 (7.03 – 7.27) | <0.0001 |
| **Lactate (mmol/L)** | 4.90 (2.4 – 8.9) | 3.1 (1.7 – 5.6) | 6.2 (3.2 – 10.0) | <0.0001 |
| **Creatinine (mmol/L)** | 108 (87.0-134.0) | 91.0 (74.0 – 111.0) | 120.0 (100.0 -152.0) | <0.0001 |

CPR=cardio-pulmonary resuscitation

### Predictors of Outcome

A set of candidate predictor variables were initially investigated for associations with the primary outcome by univariate logistic regression - results of which can be found in Table 3. In a full model, only lactate and pH were co-linear; pH was selected for use in the variable model owing to clinical relevance and a stronger statistical association. To minimise overfitting, further model refinement involved selecting variables with the strongest evidence of clinical relevance, statistical association and practical availability for application at the point of arrival to a HAC. This led to a reduced set of seven predictor variables used in the final model: age, unwitnessed arrest, initial rhythm, presence of two rhythms (defined as two out of three of VF/PEA/Asystole in a single ROSC cycle), use of any epinephrine during the cardiac arrest, pH and absent pupil reactivity (Table 4).

**Table 3:**
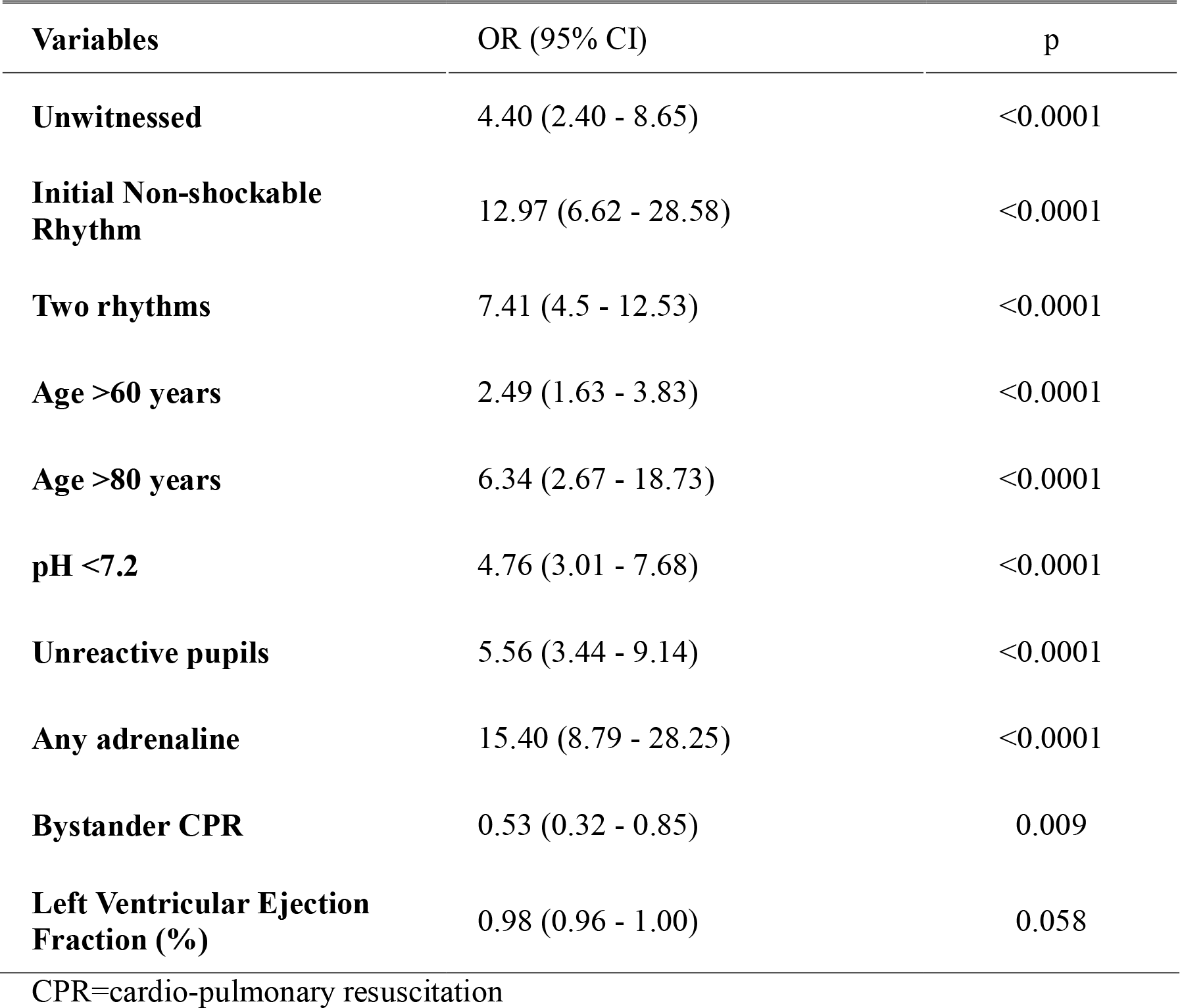
Univariate Association with the Primary End-Point

**Table 4:** Multivariable Analysis for the Primary End-Point

| **Variables** | **OR (95% CI)** | **p** | **Log (OR)** |
| --- | --- | --- | --- |
| **Unwitnessed** | 3.28 (1.42 - 7.60) | 0.006 | 1.19 |
| **Initial Rhythm** | 5.10 (2.18 - 11.96) | <0.0001 | 1.63 |
| **Unreactive pupils** | 2.57 (1.31 - 5.03) | 0.009 | 1.08 |
| **Age>60** | 2.91 (1.50 - 5.65) | 0.002 | 1.07 |
| **Age>80** | 6.95 (1.86 - 25.98) | 0.004 | 1.94 |
| **Two Rhythms** | 2.95 (1.56 - 5.59) | 0.001 | 0.79 |
| **pH<7.2** | 2.57 (1.31 - 5.03) | 0.016 | 0.94 |
| **Adrenaline** | 7.62 (3.49 - 16.63) | <0.0001 | 2.03 |

Frequency of missing variables of each predictor was deemed to be missing completely at random and so multiple imputation was considered to be an appropriate method of handling the missing predictor variable data, for which 50 imputations were performed, the results of which were pooled using Rubin’s Rule (14).

For risk score development, continuous variables in the multivariable model (age and pH) were categorised. In the multi-variate analysis, certain variables exhibited higher association with the primary outcome but, for simplicity of use in an emergency setting, each predictor was assigned only one point. However, since administration of epinephrine was a particularly strong predictor, 2 points were assigned to this variable. A value of 7.20 was used for pH and since age showed a quadratic relationship with the outcome, three age categories were used with cut-off points at 60 years (1 point) and 80 years for age (2 extra points - 3 points in total). These thresholds all gave low Akaike information criterion (AIC) model values and were deemed clinically relevant. Results from the multivariable model can be found in Table 4.

### Derivation of the two risk scores

A simplification of the above multi-variable model was used to create the final risk-score. The MIRA_2_CLE_2_ score assigns a point for each category in the model but also assigns an additional 2 points point for those over 80 years and for those on epinephrine, thus ranging from 0-10 (Figure 2). Rates of patients in each MIRA_2_CLE_2_ score are shown in Supplementary Figure 1.

**Figure 2.**
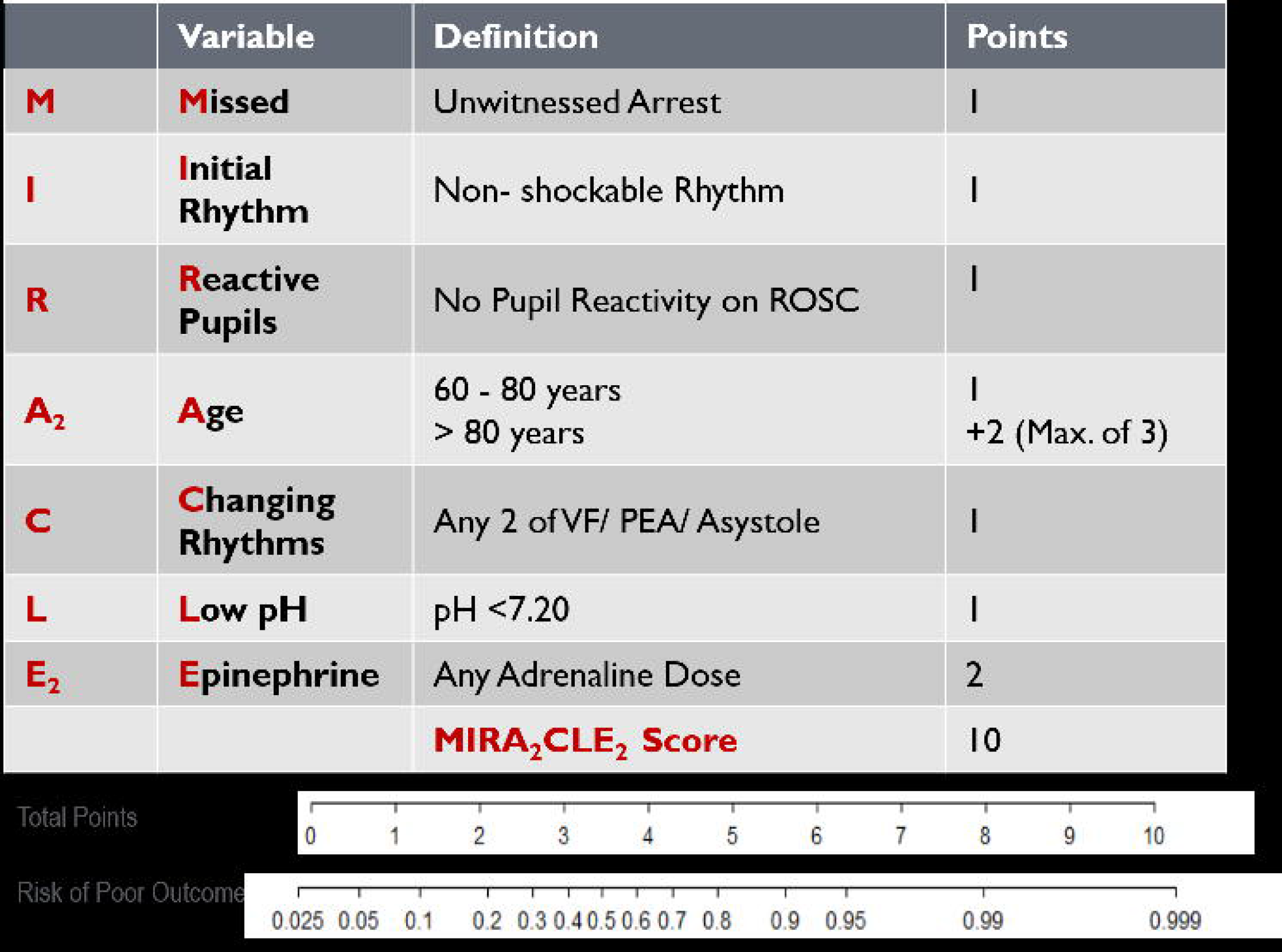
The MIRA_2_CLE_2_ score. Each variable is applied on admission and is assigned points as shown in the Figure. An extra 2 points are gained for Age >80 years (maximum of 3 if >80 years) and for epinephrine use during the cardiac arrest. A nomogram shows prediction of a poor outcome from the logistic regression model fitted to the risk score (R package rms). The risk changes in a non-linear fashion to the score and is most sensitive to changes in the score in the middle of the scale.

MIRA_2_CLE_2_had a discrimination of 0.901 AUC with internal validation median bootstrap estimate of 0.901 (95% BCa CI 0.864-0.927). MIRA_2_CLE_2_had a favourable calibration slope of 1.070 and a similar result was observed for internal validation: median bootstrap of 1.067 (95% BCa CI 0.863-1.254). The ROC curve for the MIRA_2_CLE_2_ score in the development cohort can be found in Figure 3. There was a significant and stepwise increase in the primary end-point as the MIRA_2_CLE_2_ score increased (p<0.0001) (Supplementary Figure 2 and Take Home Figure).

**Figure 3.**
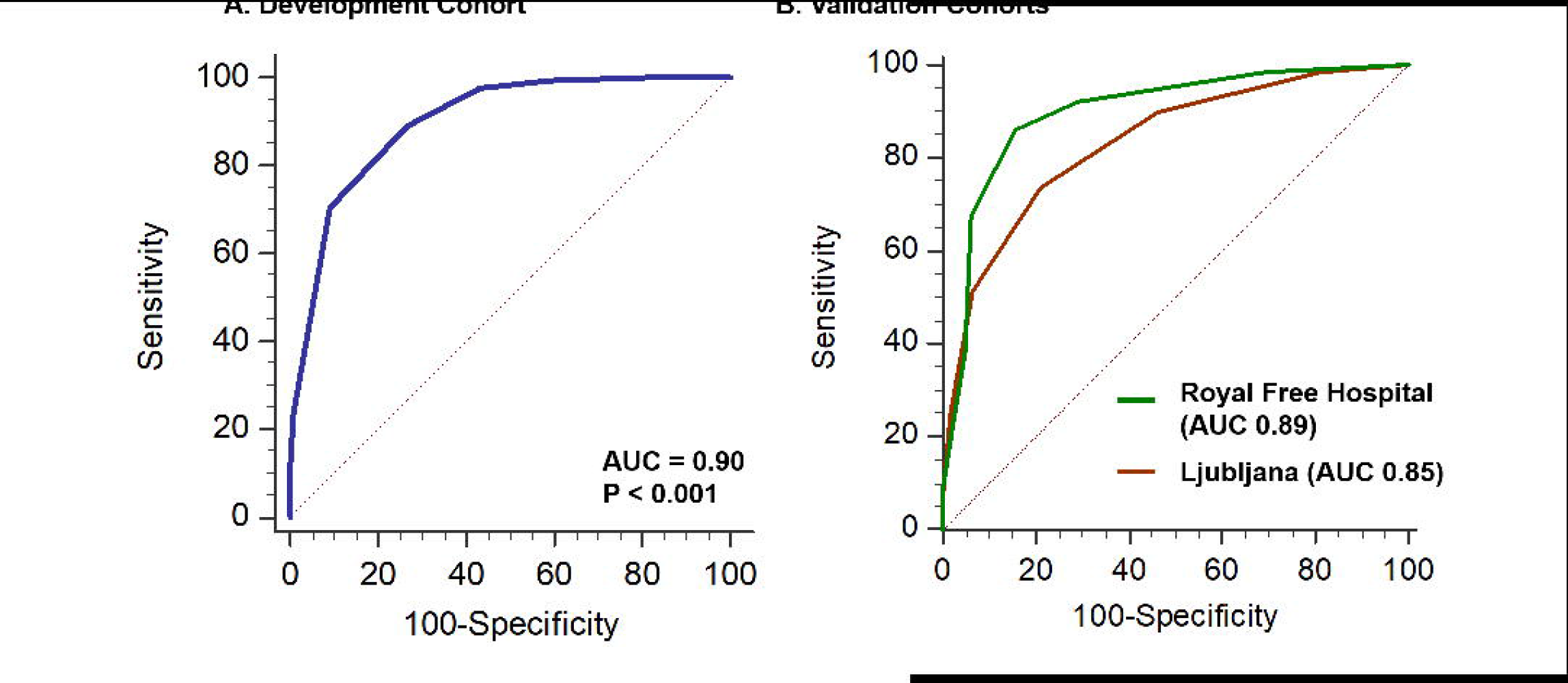
Receiver Operating Curve for the development and external validation cohorts. (Panel A) The AUC in the development cohort was 0.90 and (Panel B) was 0.85 in the Ljubljana cohort (Green) and 0.91 in Royal Free Hospital (Red). AUC=Area under the curve

### Validation Cohorts

External validation was performed in a cohort of 326 patients from the University Medical Center, Ljubljana, Slovenia admitted between January 2013 and December 2017 and 148 patients from the Royal Free Hospital (RFH), London admitted between 1^st^ January 2016 and 30^th^ March 2018 (Supplementary Table 1). All patients met inclusion and exclusion criteria and there was no overlap of patients between any cohorts. The frequency of predictor variables used in the final risk scores described above was lower in the Ljubljana and RFH cohorts than in development dataset (Supplementary Table 1). The primary end-point in the Ljubljana cohort was poor neurological outcome (CPC 3-5) at hospital discharge and in the RFH cohort was poor neurological outcome at 6 months (CPC 3-5). After imputing missing values, MIRA_2_CLE_2_ had an AUC of 0.85 in the Ljubljana validation cohort with a calibration slope of 0.897. The MIRA_2_CLE_2_ score performed more favourably in the RFH external validation cohort with an AUC of 0.91 and a calibration slope of 0.90 (Figures 3 and 4; Supplementary Figures 3 and 4).

### Discrimination Performance of the MIRA_2_CLE_2_ by Risk Groups

Three risk groups were created (**Low** - MIRA_2_CLE_2_ 0-2; **Intermediate** - MIRA_2_CLE_2_ 3-4; **High** - MIRA_2_CLE_2_≥5). From patients with complete data in the KOCAR registry, 72 patients (23.4%) were in the low risk group, 92 patients (29.9%) in the intermediate and 143 patients (46.5%) in the high-risk group. The primary end-point occurred in 5.6% of those with low-risk, 55.4% of those with intermediate-risk and 92.3% with high-risk. Assuming patients in the low risk group to be negative (and all others positive), this would give the score an estimated 97.9% sensitivity in the KOCAR population, 89.2% in the Ljubljana and 90.8% in the RFH cohorts. Alternatively, assuming patients in the high-risk group to be positive (and all others negative) this would give the score an estimated 90.8% specificity in the KOCAR population, 95.1% in the Ljubljana and 93.9% in the RFH cohorts (Supplementary Table 2).

### Discrimination Performance of the MIRA_2_CLE_2_ Score in Patients Undergoing Early Angiography

A pre-specified analysis of the performance of the MIRA_2_CLE_2_ score in patients referred for early angiography and subsequent revascularization was performed. Early angiography was performed in 262 patients from the KOCAR registry, of whom 213 had complete data. From these patients, the AUC for the primary end-point was 0.903 with a calibration slope of 1.19. 93.8% of patients with a MIRA_2_CLE_2_ score ≥5 had poor neurological outcome. In the Ljubljana external validation cohort, 223 patients had early angiography with complete data for scoring and, in this cohort, the AUC was 0.83 with a calibration slope of 0.89. 94.0% of patients with a MIRA_2_CLE_2_ score ≥5 had poor neurological outcome.

## DISCUSSION

We have derived and validated the MIRA_2_CLE_2_ risk score as a practical tool to predict poor neurological outcome at the time of index admission to a HAC. A MIRA_2_CLE_2_ score of ≥5 predicted poor neurological outcome with a specificity of 90.3% and accurately predicted poor neurological outcome in nearly half of all patients.

With increased provision of bystander CPR, early defibrillation and regionalisation of care, increasing numbers of patients are surviving OOHCA to be admitted to HACs (15). The American Heart Association (AHA) and European Society of Cardiology (ESC) both recommend emergency coronary angiography in patients with ST elevation and without ST elevation in the absence of a non-cardiac cause with evidence of ongoing ischaemia (16-18). Furthermore, there is increasing availability of MCS devices which might be used in cases of haemodynamic instability associated with OOHCA, either at specialist centres or pre-hospital in the community. However, hypoxic brain injury sustained prior to arrival is the main driver of mortality and morbidity in survivors, with concomitantly high rehabilitation costs (19). Neurological outcome can often only be reliably determined at 72 hours, and neurological risk stratification on arrival is not currently possible (20). The European Association for Percutaneous Cardiovascular Interventions (EAPCI) and AHA guidelines suggest that “favourable cardiac arrest circumstances” should be present before consideration for angiography; however this guidance is ambiguous and of limited benefit in deciding which patients should undergo invasive investigation (18, 21) Consonant with this, the recently published COACT trial demonstrated no mortality benefit of early compared to delayed angiography with outcome primarily driven by hypoxic brain injury (22).

Several tools for risk prediction have been developed but have potential limitations in the primary cardiac OOHCA cohort. The CAHP score is a normogram which was derived at the time of hospital admission with an AUC of 0.93 but is potentially more time-consuming to use in an emergency situation, only predicts short term survival (ITU discharge) so could underestimate late recovery and had poorer performance in an independent external validation cohort (AUC 0.75) (8, 9). The TTM score is also a relatively complex 10 variable multi-point score only applied at the time of ITU admission with satisfactory performance of an AUC of 0.84 but without external validation to date (9).

The MIRA_2_CLE_2_ risk score has been designed for ease of use in OOHCA patients at the point of admission to inform immediate decision making. We allocated a single point to each parameter for ease of use in an emergency setting, other than age and the use of epinephrine. Satisfactory discrimination performance in two external validation cohorts with differing rates of predictors provides robust assurance of its validity in this setting. The score predicted a low risk of poor outcome (MIRA_2_CLE_2_≤2) in over a quarter of patients while also predicting high risk (MIRA_2_CLE_2_≥5) in nearly half of patients, suggesting clinical applicability across a range of comatose patients with OOHCA. It is important to note that the intention of the score is not to replace clinical assessment but to provide an objective evaluation of neurological risk prior to a decision on delivery of potentially expensive and invasive therapies.

Several of the variables incorporated into the score, such as a witnessed arrest, initial rhythm and age are well established as being associated with outcome (23). The intra-arrest use of epinephrine in the PARAMEDIC-2 trial was not associated with improvement in post-discharge neurological recovery compared with placebo and this may represent an important surrogate of prolonged low-flow time and haemodynamic instability (24). Current guidelines continue to recommend its routine use during OOHCA, but this practice might vary in other geographic locations which could affect the discrimination value of the score in other healthcare settings (25, 26). Arterial blood gas values inputted into the score were available immediately on admission (within 30 minutes of arrival). Since it is established that resuscitative efforts might alter the values of pH, later measurement might have a significant negative impact on the performance of the score. The specificity of pupillary reflexes after ROSC in isolation for neurological outcome after OOHCA is [50% but had an odds ratio of 2.47 in our multi-variable analysis and remained a useful objective parameter in this prediction model (27). While several variables, such as zero- and low-flow times have previously been well established markers of a poor outcome, they are often unknown or inaccurately recorded at the time of admission (23), This can lead to challenges in predicting neurological outcome on arrival, especially in comatose patients treated with hypothermia. Hence, there remains significant ambiguity and limited objective guidance to support early decision making at HACs, which should have as high a specificity as possible to avoid under-treatment of cases where interval neurological recovery remains possible.

Several invasive therapies are currently available in the treatment of patients with OOHCA. Early angiography and revascularization where appropriate are currently recommended by multiple society guidelines (18, 21). The high accuracy of the MIRA_2_CLE_2_ risk score in patients treated with early angiography (21), where in this study approximately 94% of patients with a score ≥5 sustained a poor outcome, provides assurance of its validity in modern pathways of care and may, importantly, identify a specific group where an early invasive approach might be ultimately futile. Similarly, the score has substantial potential in guiding appropriate provision of MCS devices after OOHCA. There remains significant uncertainty regarding whether there is a role for MCS in OOHCA, where more robust indications for use in a centre or in the community (e-CPR) are fundamental for effective resource allocation but currently remain crude (28). Finally, methods of early neuro-prognostication after OOHCA might be useful in under-resourced medical systems or in times of crisis such a global pandemic, to avoid loss of life while also pragmatically preserving precious resources. Hence, the MIRA_2_CLE_2_ score, with further evaluation, has the potential to be incorporated into future research studies evaluating optimal treatment strategies for OOHCA by improving patient selection and excluding patients with severe neurological insult.

## Limitations

The risk score was derived and validated in retrospective cohorts, albeit with a thorough methodology and with internal and external validation. While this enabled collection of a rich dataset with a highly protocolized pathway of care, there is a risk of bias. The predictive accuracy may not be transferrable to non-HACs without access to immediate 24-hour coronary angiography, MCS, cardio-thoracic surgery and specialist intensive care expertise. The primary outcomes in one validation group differed by time-point due to availability of data and this may have affected the results, though it is established that neurological injury at discharge is usually sustained up to 1 year after discharge (29). Pupil reactivity and blood gas analysis in the Ljubljana cohort were recorded on ITU admission which might have affected the discriminant thresholds of these variables and might explain the marginally reduced performance of the score in this cohort. In contrast, the score provided higher discrimination value in the RFH validation cohort where all variables were applied directly on admission to the HAC, which may indicate that this tool should optimally be applied in this setting and provides an indication of its validity when applied at this time-point. Nonetheless, the risk score should be further prospectively validated in larger cohorts, especially immediately on arrival to HACs and across different systems of care before routine use.

## Conclusions

The MIRA_2_CLE_2_ risk score is a simple and practical tool for application on arrival a heart attack centre which can predict poor long-term neurological recovery with high accuracy.

## Data Availability

Data is not available for public distribution due to research ethics committee guidelines.

---

**Take Home Figure.**
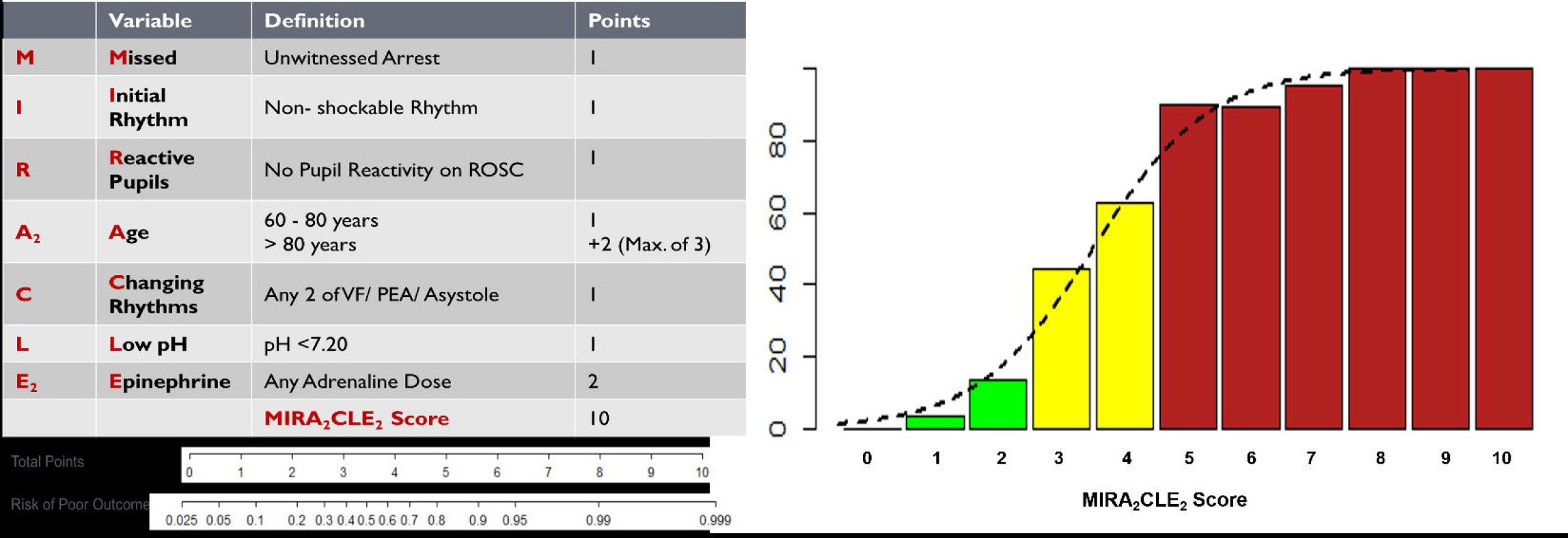
The MIRA_2_CLE_2_ score. The right-hand panel shows the observed and expected event rate (as a dotted line based on a logistic regression model of the risk score) of poor neurological outcome (CPC 3-5) at 6 months.

